# Higher mortality in male patients with Takotsubo cardiomyopathy appears to be related to higher complication rates

**DOI:** 10.1101/2025.08.13.25333624

**Authors:** Ashwin Siby, Mehrtash Hashemzadeh, Mohammad Reza Movahed

## Abstract

**Background:** Mortality in male patients with Takotsubo cardiomyopathy appears to be double than the mortality rate of Women. The goal of this study was to evaluate whether the higher mortality is related to a higher complication rate in male adults

**Methods:** Using ICD-10 codes for Takotsubo cardiomyopathy, we evaluated differences in the occurrence of complications between Men and women.

**Results:** A total of 199,890 patients had a diagnosis of Takotsubo cardiomyopathy, with 4,770 male and 165,120 female patients. All major complications are significantly higher in men than women, despite multivariate adjustment for age and cardiovascular risk factors. Cardiogenic Shock: 9.88% vs 5.98% p<0.001, OR: 1.57, 1.43-1.73, Atrial Fibrillation: 23.96% vs 20.12%, p<0.001, OR:1.55, CI 1.45-1.66, Cardiac Arrest: 5.71% vs 2.94%, p<0.001, OR: 1.71, CI 1.51-1.94, Congestive Heart Failure: 39.52% vs 35.18% p <0.001, OR: 1.23, CI:1.16-1.30, Stroke: 7.45% vs 4.94%, p<0.001, OR: 1.51, CI:1.36-1.68).

**Conclusions:** All major cardiovascular complications are higher in men compared to women with a diagnosis of Takotsubo cardiomyopathy, as a plausible explanation for the higher mortality in men.

## Introduction

Takotsubo Syndrome, also known as Takotsubo Cardiomyopathy (TC), is a stress-induced cardiomyopathy generally triggered by emotional or physical stress and is characterized by transient regional left ventricular dysfunction [1]. Nomenclature has varied, with early labels such as ‘transient cardiac ballooning’ and ‘apical ballooning syndrome.’ Some have argued that ‘stress cardiomyopathy’ more accurately reflects the full spectrum of variants beyond the classic apical phenotype [2]. The condition was first described by Dote et al in 1991 [3], and while originally thought to be a benign condition, growing evidence shows that TC is associated with significant short-term morbidity and mortality, with complication rates comparable to those observed in acute coronary syndrome [4,5]. “On echocardiography, TC shows a circumferential wall-motion abnormality with apical ballooning extending beyond a single coronary territory; coronary angiography is typically normal, helping distinguish it from ACS. [6,7,8]. Additionally, unlike ACS, Takotsubo typically shows a modest troponin rise that is disproportionately low for the extent of diffuse/circumferential wall-motion abnormality (e.g., conventional Troponin T ≤6 ng/mL or Tropinin I ≤15 ng/mL), which favors TC over ACS [9].

Epidemiological studies consistently show that TC predominantly affects women, particularly postmenopausal women [11,12]. Emotional stressors - such as bereavement or personal trauma- are common triggers in women, whereas men more frequently develop TC following physical stressors like surgery, infection, or acute illness [4,13]. These differences suggest potential sex-based physiological mechanisms underlying TC susceptibility. Notably, anatomic variants exist; the reverse (basal) pattern tends to occur in younger patients and is almost uniformly precipitated by an identifiable stressor [14]. Though not completely understood, proposed mechanisms include catecholamine-induced myocardial stunning, coronary microvascular dysfunction, and transient left ventricular outflow tract obstruction [15,16]. This aligns with stimulant-associated reverse TC, supporting a catecholamine-excess mechanism in younger presentations. [17]. The cardioprotective effects of estrogen, including enhanced endothelial cell function and attenuated sympathetic responses, are thought to contribute to the lower severity and better outcomes typically observed in women [18].

Although less frequently diagnosed in men, studies consistently show that male patients experience worse clinical outcomes when diagnosed with TC [13,19,20]. Recent meta-analyses and registry-based studies report that men with TC have significantly higher rates of in-hospital mortality, cardiogenic shock, cardiac arrest, and other serious complications [13,19]. For example, Abusnina et al demonstrated that men with TC have more than double the risk of in-hospital death and cardiogenic shock compared to women [19]. Similarly, Maskoun et al found that men are more likely to present with severe hemodynamic instability requiring mechanical ventilation and circulatory support during hospital course [13]. These findings consistently identify male sex as an independent predictor of adverse outcomes in TC [13,19]. A recent study documented double mortality in men vs women in patients admitted with Takotsubo cardiomyopathy. However, the cause of these differences remained elusive (20).

To evaluate the reason behind the higher mortality of men suffering from TC cardiomyopathy, we hypothesized that men may suffer from a higher complication rate. Therefore, we aimed to evaluate whether male patients with TC experience higher rates of in-hospital cardiovascular complications compared to female patients by utilizing the Nationwide Inpatient Sample (NIS) database from 2016 to 2020.

## Methods

### Data Source

This study is deemed institutional review board exempt as the NIS is a publicly available deidentified database. The NIS database includes weighted discharge information for about 35 million patients each year, 20% of all inpatient admissions to nonfederal hospitals in the United States.

### Study Population

In this retrospective observational cohort study, all patients aged >18years from the 2016 to 2020 NIS database were included. Our target population was patients hospitalized with TC, which was identified using International Classification of Diseases, Tenth Revision (ICD-10) code I51.81.

The key complications of interest including myocardial rupture (I23.2, I23.3, I23.4, I23.5), cardiogenic shock (R57.0), atrial fibrillation(I48), cardiac arrest (I46), congestive heart failure (CHF; I52, I53, I54), and stroke (I60, I61, I62, I63) were also recorded. Prior cardiovascular risk factors, including diabetes, hypertension, hyperlipidemia, and chronic kidney disease, were evaluated. Patient demographics include age, sex, race or ethnicity, and hospital demographics, including median household income, expected primary payer, hospital bed size, location and teaching status of hospital, hospital region, control of hospital, along with average hospital length of stay, and average total charges were evaluated.

### Study Outcomes

The patient outcome examined was in-hospital complications, including cardiogenic shock, atrial fibrillation, cardiac arrest, congestive heart failure, and stroke, with diagnoses of TC. In multivariate analysis, adjustments were made for demographic factors (age, race, primary payer, median household income by zip code), hospital characteristics (bed size, teaching status, and region), and patient comorbidities (hypertension, diabetes, obesity, chronic kidney disease, chronic obstructive pulmonary disease, and dyslipidemia).

### Statistical Analysis

Patient demographic, clinical, and hospital characteristics are reported as percentages in Table 1. Odds ratios (ORs) and 95% CIs are calculated for continuous variables and proportions and 95% CIs for categorical variables. We studied outcome data over the 5-year period (2016-2020). Categorical outcomes were assessed using chi-squared analysis. Multivariate logistic regression models were constructed to evaluate the association between sex and the occurrence of in-hospital cardiovascular complications, including cardiogenic shock, atrial fibrillation, cardiac arrest, congestive heart failure, and stroke. All analyses accounted for the NIS sampling design using appropriate survey weights to generate nationally representative estimates. Statistical significance was defined as a two-sided p-value <0.05. Statistical analyses were performed using Stata version 16.0 (StataCorp, College Station, TX).

**Table 1.**
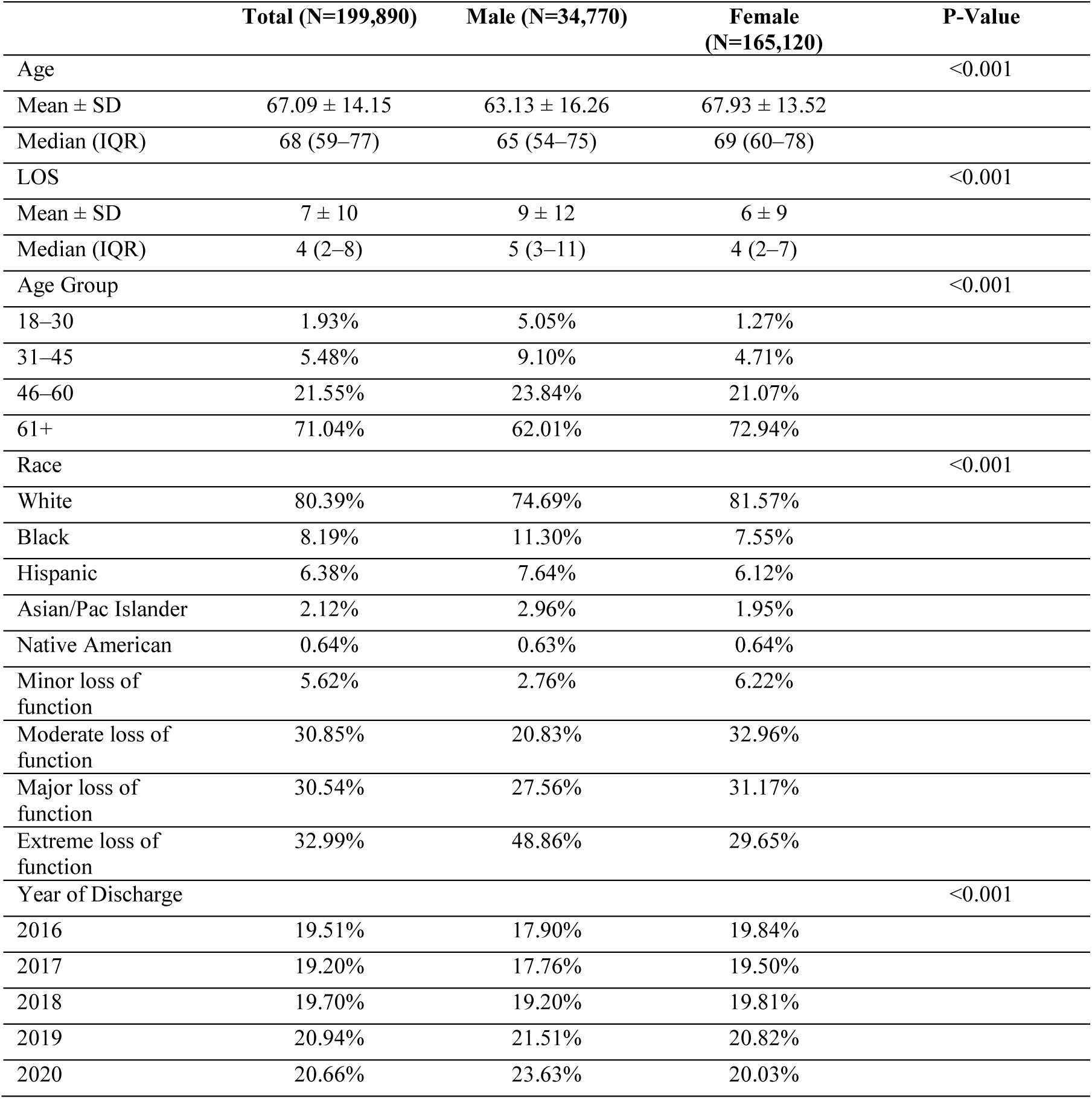
Baseline Characteristics of Patients Hospitalized with Takotsubo Cardiomyopathy, 2016–2020.

|  | **Total (N=199,890)** | **Male (N=34,770)** | **Female (N=165,120)** | **P-Value** |
| --- | --- | --- | --- | --- |
| Age |  |  |  | <0.001 |
| Mean ± SD | 67.09 ± 14.15 | 63.13 ± 16.26 | 67.93 ± 13.52 |  |
| Median (IQR) | 68 (59–77) | 65 (54–75) | 69 (60–78) |  |
| LOS |  |  |  | <0.001 |
| Mean ± SD | 7 ± 10 | 9 ± 12 | 6 ± 9 |  |
| Median (IQR) | 4 (2–8) | 5 (3–11) | 4 (2–7) |  |
| Age Group |  |  |  | <0.001 |
| 18–30 | 1.93% | 5.05% | 1.27% |  |
| 31–45 | 5.48% | 9.10% | 4.71% |  |
| 46–60 | 21.55% | 23.84% | 21.07% |  |
| 61+ | 71.04% | 62.01% | 72.94% |  |
| Race |  |  |  | <0.001 |
| White | 80.39% | 74.69% | 81.57% |  |
| Black | 8.19% | 11.30% | 7.55% |  |
| Hispanic | 6.38% | 7.64% | 6.12% |  |
| Asian/Pac Islander | 2.12% | 2.96% | 1.95% |  |
| Native American | 0.64% | 0.63% | 0.64% |  |
| Minor loss of function | 5.62% | 2.76% | 6.22% |  |
| Moderate loss of function | 30.85% | 20.83% | 32.96% |  |
| Major loss of function | 30.54% | 27.56% | 31.17% |  |
| Extreme loss of function | 32.99% | 48.86% | 29.65% |  |
| Year of Discharge |  |  |  | <0.001 |
| 2016 | 19.51% | 17.90% | 19.84% |  |
| 2017 | 19.20% | 17.76% | 19.50% |  |
| 2018 | 19.70% | 19.20% | 19.81% |  |
| 2019 | 20.94% | 21.51% | 20.82% |  |
| 2020 | 20.66% | 23.63% | 20.03% |  |

### Results

In this analysis of the Nationwide Inpatient Sample (NIS) from 2016 to 2020, we identified a weighted total of 199,890 adult patients hospitalized with a primary diagnosis of TC using the ICD-10 codes. The cohort included 34,770 male patients and 165,120 female patients. All analyses were conducted following the application of population discharge weights to provide nationally representative estimates.

### Baseline Characteristics

The baseline characteristics of the study population are presented in Table 1. The mean age of the overall cohort was 67.09 ± 14.15 years, with male patients being significantly younger than female patients (63.13 ± 16.26 years vs 67.93 ± 13.52 years; *P* < 0.001). Men also had a longer median length of hospital stay compared to women (5 [IQR 3–11] days vs 4 [IQR 2–7] days; *P* < 0.001).

In terms of age distribution, male patients were more likely to be younger, with 5.05% aged 18– 30 years compared to 1.27% of female patients. Female patients were more likely to be aged 61 years or older (72.94% vs 62.01%; *P* < 0.001). There were significant differences in racial distribution, with Black and Hispanic patients comprising a larger proportion of the male group.

Male patients were more likely to have Medicaid (14.68% vs 9.96%) and self-pay hospitalizations (4.28% vs 2.34%), while female patients were more likely to be covered by Medicare (65.29% vs 53.98%; *P* < 0.001). Most hospitalizations occurred in large, urban teaching hospitals across all groups.

### In-Hospital Complications

Compared to female patients, men had a higher prevalence of smoking (28.75% vs 27.13%; *P* = 0.008) and diabetes (25.61% vs 23.81%; *P* = 0.002). Conversely, women had a significantly higher prevalence of hypertension (67.82% vs 61.52%; *P* < 0.001) and hyperlipidemia (47.13% vs 38.86%; *P* < 0.001). Chronic kidney disease (CKD) was more common in male patients (18.39% vs 14.84%; *P* < 0.001). These findings are summarized in Table 2.

**Table 2.**
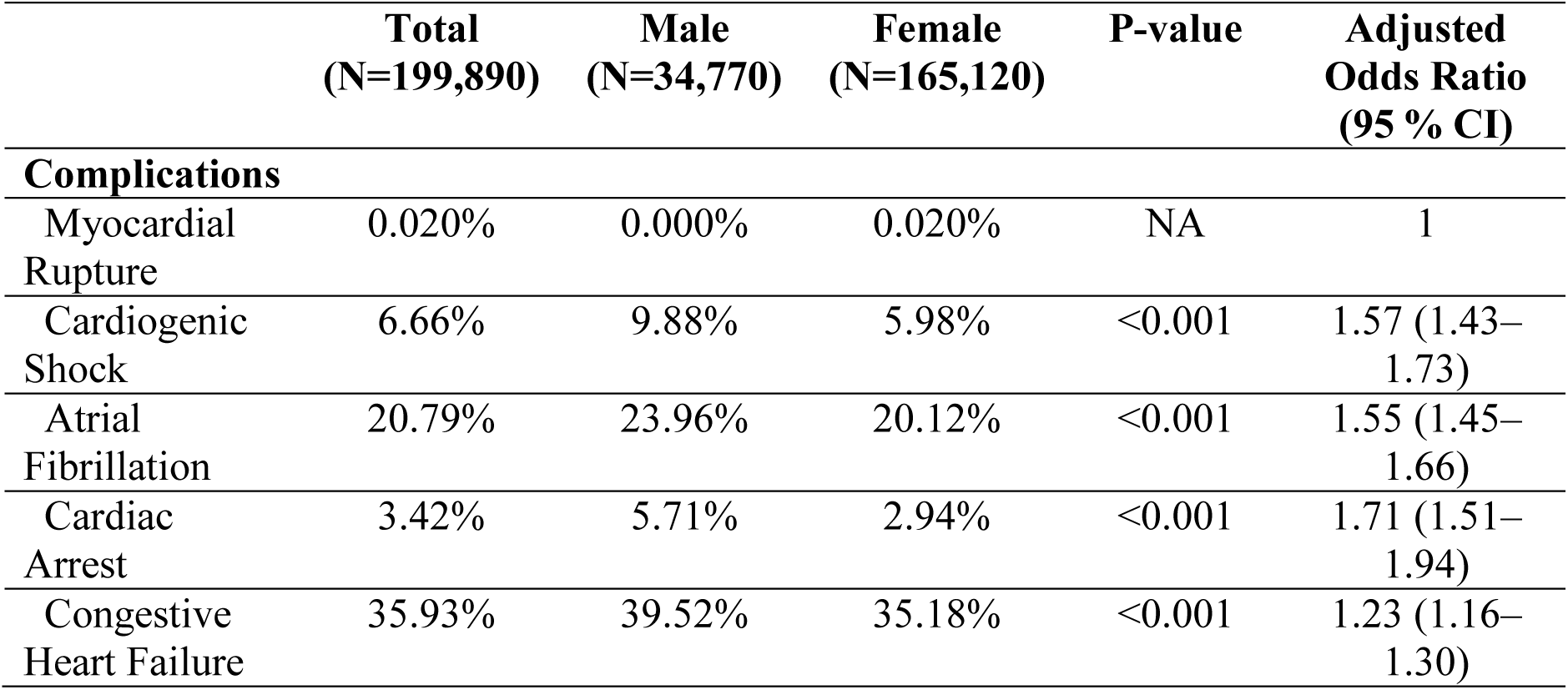
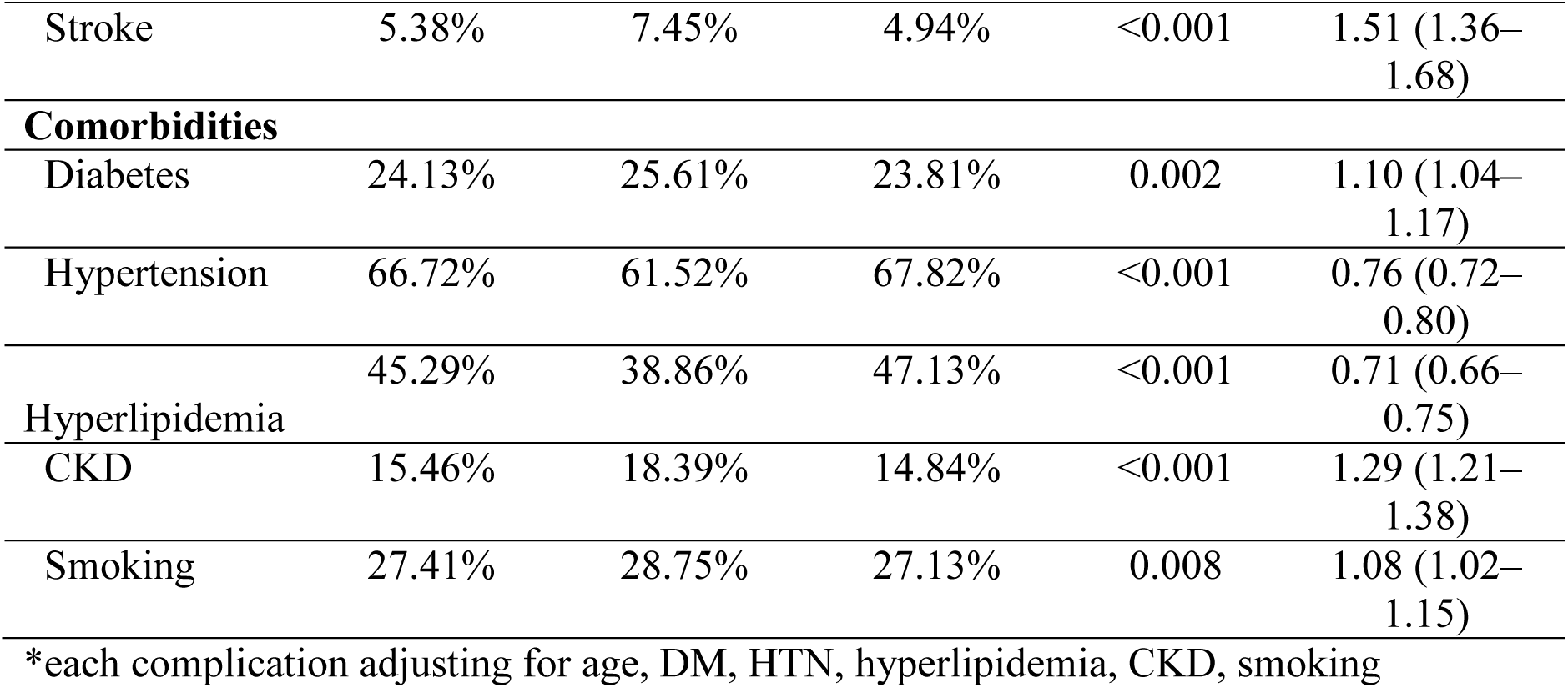
In-hospital Complications and Baseline Comorbidities by Sex.

Male patients also had significantly higher rates of cardiogenic shock (9.88% vs 5.98%; OR 1.72; 95% CI, 1.57–1.89), atrial fibrillation (23.96% vs 20.12%; OR 1.25; 95% CI, 1.18–1.33), cardiac arrest (5.71% vs 2.94%; OR 2.00; 95% CI, 1.77–2.25), congestive heart failure (39.52% vs 35.18%; OR 1.20; 95% CI, 1.14–1.27), and stroke (7.45% vs 4.94%; OR 1.55; 95% CI, 1.40–1.72), all *P* < 0.001. The percentages of complication rates are displayed in Figure 1.

**Figure 1.**
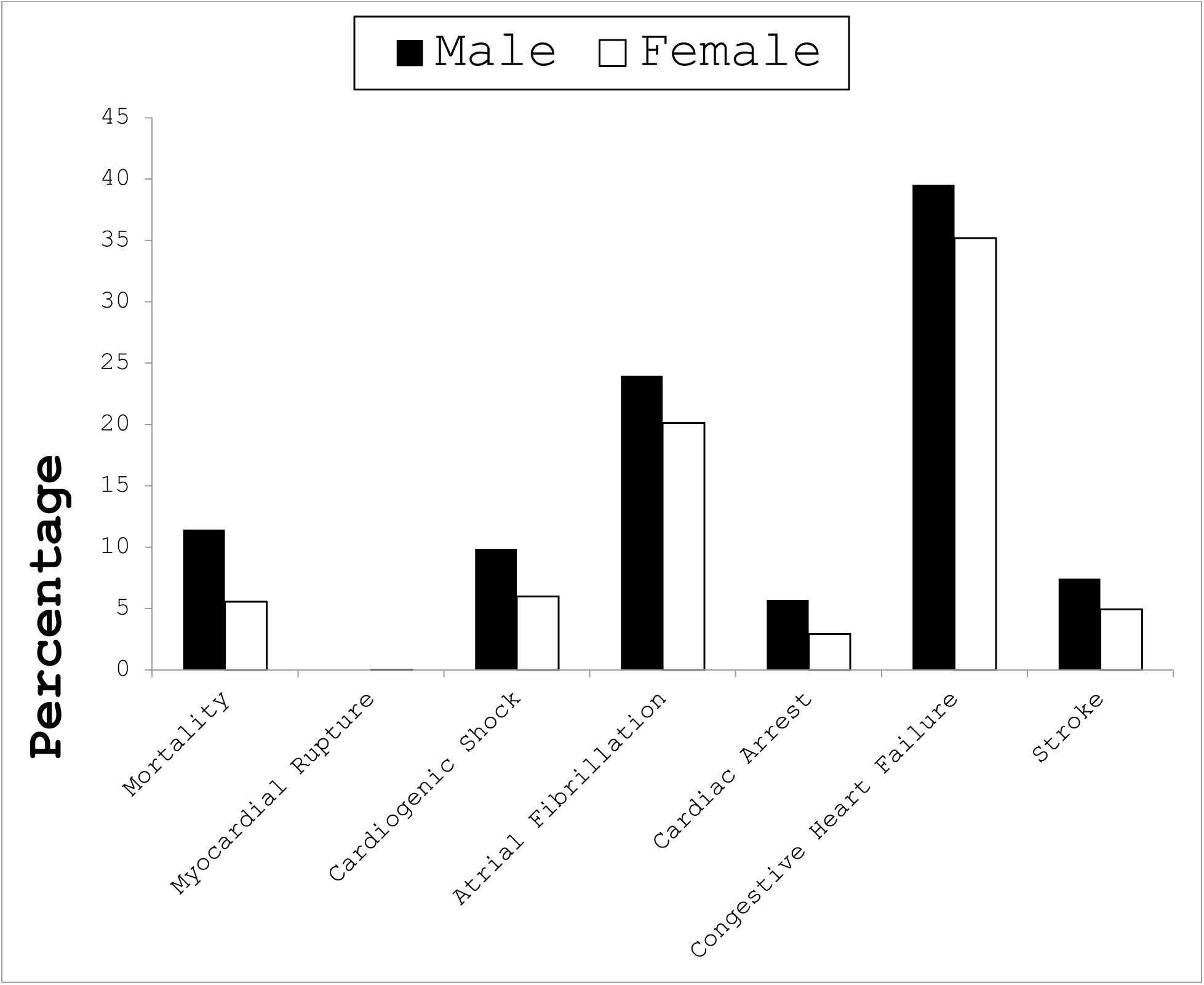
In-hospital complication rates among male and female patients with Takotsubo cardiomyopathy. Complications of Takotsubo Cardiomyopathy based on Sex.

The odds ratios for these in-hospital complications are displayed in Figure 2 as a forest plot, demonstrating that male sex was independently associated with significantly higher odds of mortality and all major cardiovascular complications.

**Figure 2.**
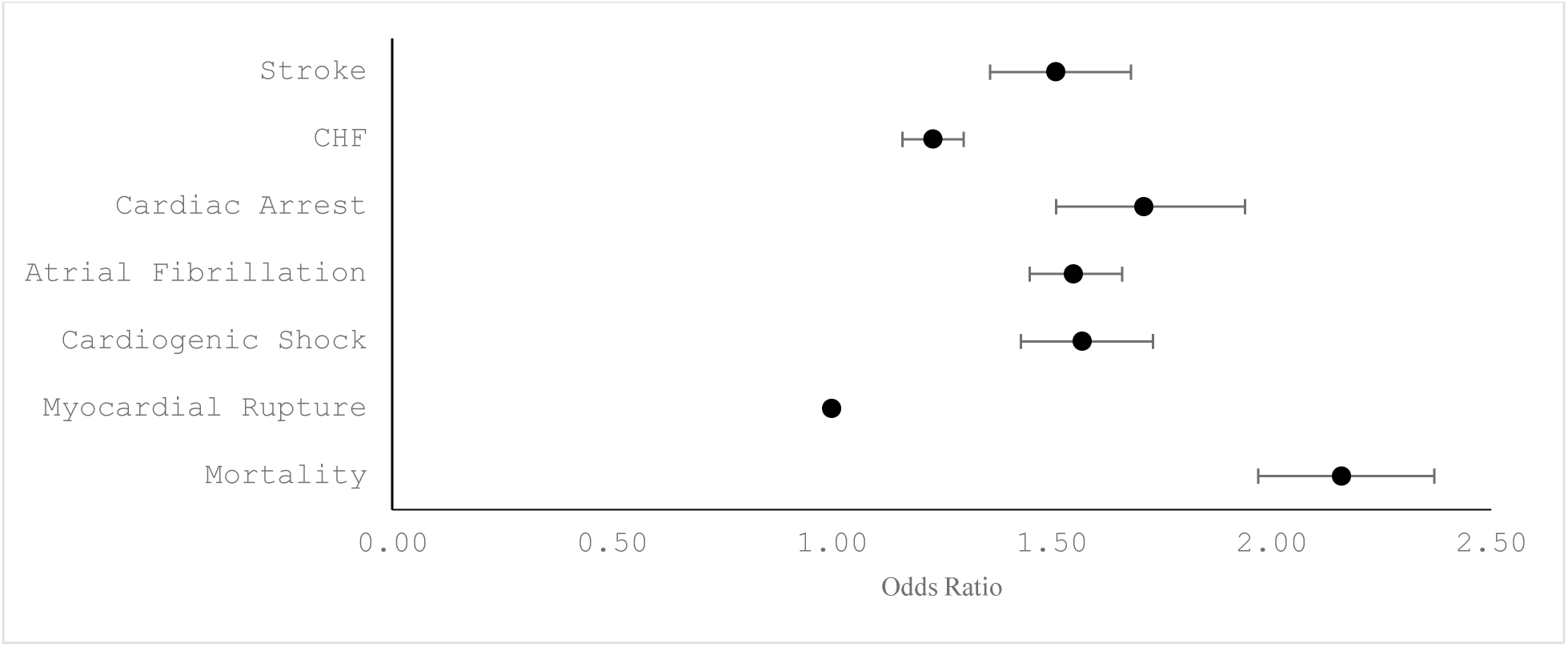
Adjusted odds ratio and 95% confidence intervals comparing male vs female patients for major in-hospital complications. Odds ratios above 1 indicate higher complication rates in men.

### Severity of Illness and Risk of Mortality

Male patients were more likely to be classified in higher All Patient Refined Diagnosis-Related Groups (APR-DRG) severity of illness and risk of mortality subclasses. Nearly half of male patients (47.68%) were categorized as having an extreme likelihood of dying compared to 29.33% of female patients (*P* < 0.001). Similarly, 48.86% of male patients were classified as having extreme loss of function, compared to 29.65% of female patients (*P* < 0.001).

### Yearly Trends

The annual incidence of TC showed a slight overall increase from 2016 to 2020, but no consistent year-over-year trend was observed. The distribution of TC hospitalizations by year and sex is shown in Table 1.

## Discussion

### Summary of Key Findings

Our analysis of a large nationwide cohort (2016-2020) demonstrates that male patients with Takotsubo cardiomyopathy (TC) experience significant worse in-hospital outcomes than female patients. Men comprised only ∼17% of TC hospitalizations yet had more than double the in-hospital mortality rate of women (approximately 11.2% vs 5.5%) [20]. They also suffered higher rates of acute complications, including cardiogenic shock, atrial fibrillation, cardiac arrest, acute congestive heart failure, and stroke during the index admission. Notably, overall in-hospital mortality for TC was high (∼6.5%) and showed no significant improvement from 2016 to 2020 [20]. These findings highlight a pronounced sex disparity: although TC predominantly affects women, men who develop TC tend to have a markedly more severe clinical course and a higher risk of death. The excess complications in men appear to be a major contributor to their elevated mortality. In summary, male sex emerged as a clear risk factor for adverse outcomes in TC in this contemporary national dataset.

These observations align with prior work. A recent meta-analysis of twelve cohorts found that men carried roughly double the odds of in-hospital death compared with women [19].

Additionally, in both unmatched and propensity-matched cohorts, Arcari et al. found that male sex was independently associated with significantly higher rates of cardiogenic shock, in-hospital mortality, and long-term mortality among patients with Takotsubo syndrome [21]. A Veterans Affairs cohort of 641 patients showed in-hospital mortality of 8.1% in men vs 1.0% in woman [22]. Using a U.S nationwide sample from 2011-2108, Vincent et al. found that male sex conferred an adjusted odds ratio of 2.4 for mortality as well as higher rates of shock and arrhythmias [23]

### Comparison with Existing Literature

These observations align with prior work. A recent meta-analysis of twelve observational cohorts found that men carried roughly double the odds of in-hospital death compared with women (RR 2.17, 95% CI 1.77–2.67) and had a significantly higher risk of cardiogenic shock (RR 1.66, 95% CI 1.29–2.12) [10]. The consistency of these findings across a large, pooled population supports the idea that male sex is an independent predictor of worse outcomes in Takotsubo syndrome, even after accounting for differences in age and baseline comorbidities.

In support of this, Arcari et al. analyzed both unmatched and propensity-matched cohorts from the GEIST registry and similarly found that male sex was associated with significantly higher rates of in-hospital mortality, cardiogenic shock, and long-term mortality. Even after matching men and women on clinical characteristics and triggers, the mortality gap persisted, with men experiencing in-hospital mortality of 8% versus 3% in women, and cardiogenic shock occurring in 16% of men versus 6% of women. [21]. These rates are remarkably consistent with what we observed in our cohort, further reinforcing that the male disadvantage in TC outcomes is not a product of selection bias or differences in presentation alone.

Additional evidence comes from a Veterans Affairs cohort study of 641 patients, which found in-hospital mortality rates of 8.1% in men compared to only 1.0% in women [22]. While the study population was predominantly male, the mortality difference was striking and held up even after adjustment for baseline differences, indicating that sex itself likely plays a role in determining prognosis. This is notable because it supports the idea that even within a more homogenous health system, the outcome disparity remains.

Further confirmation comes from Vincent et al., who used a large U.S. nationwide sample from 2011 to 2018 and found that male sex conferred an adjusted odds ratio of 2.4 for in-hospital mortality. They also reported higher rates of cardiogenic shock, ventricular arrhythmias, and respiratory failure in men [23]. These findings are consistent with both the direction and magnitude of the associations seen in our study, suggesting that the pattern has remained stable over time despite advances in supportive care. In combination, these prior studies and our current findings point to a persistent and reproducible difference in outcomes based on sex, with male patients consistently demonstrating a higher risk for acute complications and death following Takotsubo cardiomyopathy.

### Potential Mechanisms

The reasons behind worse outcomes in men with Takotsubo cardiomyopathy are likely multifactorial. One proposed explanation is the absence of estrogen-mediated cardio protection. Estrogen is thought to blunt the effects of catecholamine surges, improve endothelial function, and modulate sympathetic activation—mechanisms that may help limit myocardial injury during acute stress [24]. Preclinical studies have shown that activation of estrogen receptors can reduce the extent of myocardial stunning and apical ballooning in stress models [25]

Triggering events also differ by sex. Men are more likely to develop TC in response to physical stressors such as infection, surgery, or trauma, while women more often present after emotional stressors [4,13]. The physical stress subtype is associated with worse outcomes and more frequent ICU-level complications. Additionally, men in our cohort had a higher prevalence of comorbidities such as chronic kidney disease and diabetes, which could contribute to more severe presentations and reduced cardiac reserve. These clinical and physiological differences likely contribute to the higher observed rates of cardiogenic shock, arrhythmias, and mortality in male patients.

### Clinical Implications

Recognizing the sex-based differences in TC outcomes has important clinical implications for patient management and risk stratification. Male patients with Takotsubo syndrome should be considered high-risk and may warrant more intensive monitoring and supportive care during their acute illness. For instance, our findings suggest that a middle-aged or elderly man diagnosed with TC is much more likely to develop cardiogenic shock, arrhythmias, or respiratory failure than a female counterpart. Practically, clinicians should maintain a lower threshold to manage male TC patients in an intensive care unit setting, where close hemodynamic monitoring and rapid intervention (vasopressors, mechanical circulatory support, ventilatory support) can be provided. In a prior study, men with TC frequently required invasive ventilation and vasopressor therapy at significantly higher rates [22]. Anticipating this need can facilitate early placement of arterial lines, consideration of intra-aortic balloon pump or Impella in cases of evolving shock, and consultation with heart failure/cardiomyopathy specialists. In short, male sex can be viewed as a potential red flag for ICU-level care in Takotsubo cardiomyopathy.

Another implication is in diagnostic vigilance. Because most TC patients are female, clinicians might not immediately recognize Takotsubo syndrome in a male patient, potentially attributing cardiac dysfunction to ischemic cardiomyopathy or other causes. Our findings reinforce that although less common, TC does occur in men, and when it does, it is often under dire circumstances. Prompt diagnosis is important because it can focus management on supportive care and alert providers to expect complications. The literature speculates that the higher mortality in men could partly stem from delayed recognition or treatment disparities [19].

Addressing this requires education and awareness: cardiologists and intensivists should be aware that the so-called “broken heart syndrome” is not exclusive to women, and its presentation in men often signals a grave clinical scenario that demands aggressive support.

In terms of a prognostic standpoint, sex should be incorporated into risk assessment models for Takotsubo. Our data, in conjunction with previous findings, show male sex is an independent predictor of in-hospital mortality. Risk scores or clinical decision tools for TC (whether for predicting complications or guiding triage) would likely be improved by including male gender as a key variable. For example, a hypothetical TC risk stratification might assign a higher score to men, prompting earlier transfer to tertiary centers or consideration of advanced therapies.

Clinicians should also counsel patients and families that TC is not always benign – especially in male patients, who face an elevated risk of acute decompensation. This is important for shared decision-making and setting appropriate expectations for monitoring and possible interventions (such as temporary mechanical support or ICU therapies).

## Limitations

Several limitations of our study should be acknowledged when interpreting the results. First, this was an observational analysis of an administrative database (the Nationwide Inpatient Sample), which relies on ICD coding for identifying Takotsubo cardiomyopathy and complications.

Misclassification is possible; some cases of TC might have been missed or conversely, coding errors could include miscoded cases. However, prior validations suggest ICD codes for stress cardiomyopathy are reasonably specific. Additionally, the NIS database lacks granular clinical detail, as we did not have data on the precipitating stressor for each case, the exact timing of complication onset, laboratory values (e.g. catecholamine levels, troponin), or imaging findings beyond what can be inferred from codes. Thus, our ability to explore the mechanism was limited to inference from diagnosis codes and demographics. We could not directly confirm, for example, which patients had “secondary” TC due to an underlying illness vs “primary” TC, nor assess differences in TC anatomical subtypes (apical vs basal ballooning) between sexes. Finally, unmeasured confounders may influence our results. We adjusted for available comorbidities, but factors like severity of illness scores, treatment differences, or socioenvironmental stress factors were not captured. It is possible that men received different management or presented later than women, which we could not ascertain but could contribute to outcome differences.

Another limitation is that our endpoint was in-hospital outcomes only. We did not track patients after discharge, so long-term mortality or recurrence rates by sex could not be evaluated in this dataset. Prior studies indicate men may also have worse long-term outcomes [21] but our analysis cannot address that. Additionally, our data represent U.S. hospitalizations; the findings may not be fully generalizable to other countries or healthcare systems. Cultural differences in stress exposure, healthcare-seeking behavior, or treatment protocols might influence sex-based outcomes. Lastly, while the study period ending in 2020 provided up-to-date insights, it also coincided with the beginning of the COVID-19 pandemic. We did not specifically analyze the impact of COVID-related stress or myocardial injury on TC cases, an area that could be explored further, as acute infections (including COVID-19) have been reported to trigger Takotsubo syndrome [27]. Despite these limitations, the study’s large sample size and congruence with existing literature lend credence to our conclusions. We believe the core finding that male TC patients have higher complication and mortality rates is robust, though the exact magnitude of risk may vary with patient populations and management differences.

## Future Directions

Our work and prior studies raise several questions for future research and opportunities to improve care. At the clinical level, there is a need to develop better risk stratification tools that account for sex, comorbid conditions, and the nature of the trigger event. Male patients presenting with TC in the context of infection, trauma, or post-operative stress likely represent a distinct high-risk phenotype that may benefit from early ICU-level care, more aggressive hemodynamic monitoring, and consideration for temporary mechanical support in the setting of shock. Whether tailored management pathways for these patients improve outcomes remains an open question and should be studied prospectively.

Long-term outcomes also deserve further investigation. Several studies, including registry data, suggest that male patients may not only have higher in-hospital mortality but also worse long-term survival, potentially related to non-cardiac comorbidities or late cardiovascular complications [21,22,23].. Studies with extended follow-up could help determine whether male survivors of TC would benefit from more intensive outpatient monitoring, heart failure therapy, or arrhythmia surveillance.

Finally, awareness of the prognostic importance of sex in Takotsubo cardiomyopathy should be integrated into educational efforts and possibly even into clinical guidelines. Despite being historically viewed as a condition predominantly affecting older women, male patients clearly represent a higher-risk subgroup and should not be overlooked or assumed to follow the same benign course. Future updates to consensus statements or position papers on stress cardiomyopathy should consider highlighting these sex-specific risks and management considerations.

## Conclusion

In this large, nationally representative cohort of patients hospitalized with Takotsubo cardiomyopathy, male sex was independently associated with significantly higher rates of major cardiovascular complications, including cardiogenic shock, atrial fibrillation, cardiac arrest, congestive heart failure, and stroke. These differences persisted after adjustment for age, comorbidities, and hospital characteristics, and are consistent with findings from prior registries and meta-analyses. These higher complication rates can explain the reason behind double mortality from TC in men compared with women and underscore the need for heightened clinical vigilance, tailored risk stratification, and further research into sex-specific mechanisms and management strategies.

## Data Availability

NIS data is publicialy available

## References

1. Assad J, Femia G, Pender P, Badie T, Rajaratnam R. Takotsubo Syndrome: A Review of Presentation, Diagnosis and Management. Clin Med Insights Cardiol. 2022;16:11795468211065782. Published 2022 Jan 4. doi:10.1177/11795468211065782

2. Movahed MR. Transient cardiac ballooning is not best nomenclature for Takotsubo cardiomyopathy as it does not capture all variants of this syndrome. Stress cardiomyopathy is a much better term for this syndrome. Clin Cardiol. 2010;33(4):241–242. doi:10.1002/clc.20742

3. Dote K, Sato H, Tateishi H, Uchida T, Ishihara M. J Cardiol. 1991;21(2):203–214.

4. Templin C, Ghadri JR, Diekmann J, et al. Clinical Features and Outcomes of Takotsubo (Stress) Cardiomyopathy. N Engl J Med. 2015;373(10):929–938. doi:10.1056/NEJMoa1406761

5. Vallabhajosyula S, Barsness GW, Herrmann J, Anavekar NS, Gulati R, Prasad A. Comparison of Complications and In-Hospital Mortality in Takotsubo (Apical Ballooning/Stress) Cardiomyopathy Versus Acute Myocardial Infarction. Am J Cardiol. 2020;132:29–35. doi:10.1016/j.amjcard.2020.07.015

6. Movahed MR. Important echocardiographic features of takotsubo or stress-induced cardiomyopathy that can aid early diagnosis. JACC Cardiovasc Imaging. 2010;3(11):1200–1201. doi:10.1016/j.jcmg.2010.08.015

7. Movahed MR, Donohue D. Review: transient left ventricular apical ballooning, broken heart syndrome, ampulla cardiomyopathy, atypical apical ballooning, or Tako-Tsubo cardiomyopathy. Cardiovasc Revasc Med. 2007;8(4):289–292. doi:10.1016/j.carrev.2007.02.001

8. Donohue D, Ahsan C, Sanaei-Ardekani M, Movahed MR. Early diagnosis of stress-induced apical ballooning syndrome based on classic echocardiographic findings and correlation with cardiac catheterization. J Am Soc Echocardiogr. 2005;18(12):1423. doi:10.1016/j.echo.2005.05.017

9. Ramaraj R, Sorrell VL, Movahed MR. Levels of troponin release can aid in the early exclusion of stress-induced (takotsubo) cardiomyopathy. Exp Clin Cardiol. 2009;14(1):6–8.

10. Donohue D, Movahed MR. Clinical characteristics, demographics and prognosis of transient left ventricular apical ballooning syndrome. Heart Fail Rev. 2005;10(4):311–316. doi:10.1007/s10741-005-8555-8

11. Amin HZ, Amin LZ, Pradipta A. Takotsubo Cardiomyopathy: A Brief Review. J Med Life. 2020;13(1):3–7. doi:10.25122/jml-2018-0067

12. Donohue D, Movahed MR. Clinical characteristics, demographics and prognosis of transient left ventricular apical ballooning syndrome. Heart Fail Rev. 2005;10(4):311–316. doi:10.1007/s10741-005-8555-8

13. Maskoun W, Alqam B, Habash F, Gheith Z, Sawada SG, Vallurupalli S. Sex Differences in Stress-Induced (Takotsubo) Cardiomyopathy. CJC Open. 2022;5(2):120–127. Published 2022 Nov 18. doi:10.1016/j.cjco.2022.11.012

14. Ramaraj R, Movahed MR. Reverse or inverted takotsubo cardiomyopathy (reverse left ventricular apical ballooning syndrome) presents at a younger age compared with the mid or apical variant and is always associated with triggering stress. Congest Heart Fail. 2010;16(6):284–286. doi:10.1111/j.1751-7133.2010.00188.x

15. Prasad A, Lerman A, Rihal CS. Apical ballooning syndrome (Tako-Tsubo or stress cardiomyopathy): a mimic of acute myocardial infarction. Am Heart J. 2008;155(3):408–417. doi:10.1016/j.ahj.2007.11.008

16. Lyon AR, Bossone E, Schneider B, et al. Current state of knowledge on Takotsubo syndrome: a Position Statement from the Taskforce on Takotsubo Syndrome of the Heart Failure Association of the European Society of Cardiology. Eur J Heart Fail. 2016;18(1):8–27. doi:10.1002/ejhf.424

17. Movahed MR, Donohue D. Review: transient left ventricular apical ballooning, broken heart syndrome, ampulla cardiomyopathy, atypical apical ballooning, or Tako-Tsubo cardiomyopathy. Cardiovasc Revasc Med. 2007;8(4):289–292. doi:10.1016/j.carrev.2007.02.001

18. Ueyama T, Hano T, Kasamatsu K, Yamamoto K, Tsuruo Y, Nishio I. Estrogen attenuates the emotional stress-induced cardiac responses in the animal model of Tako-tsubo (Ampulla) cardiomyopathy. J Cardiovasc Pharmacol. 2003;42 Suppl 1:S117–S119. doi:10.1097/00005344-200312001-00024

19. Abusnina W, Elhouderi E, Walters RW, et al. Sex Differences in the Clinical Outcomes of Patients With Takotsubo Stress Cardiomyopathy: A Meta-Analysis of Observational Studies. Am J Cardiol. 2024;211:316–325. doi:10.1016/j.amjcard.2023.10.066

20. Movahed MR, Javanmardi E, Hashemzadeh M. High Mortality and Complications in Patients Admitted With Takotsubo Cardiomyopathy With More Than Double Mortality in Men Without Improvement in Outcome Over the Years. J Am Heart Assoc. 2025;14(10):e037219. doi:10.1161/JAHA.124.037219

21. Arcari L, Núñez Gil IJ, Stiermaier T, et al. Gender Differences in Takotsubo Syndrome. J Am Coll Cardiol. 2022;79(21):2085–2093. doi:10.1016/j.jacc.2022.03.366

22. Maskoun W, Alqam B, Habash F, Gheith Z, Sawada SG, Vallurupalli S. Sex Differences in Stress-Induced (Takotsubo) Cardiomyopathy. CJC Open. 2022;5(2):120–127. Published 2022 Nov 18. doi:10.1016/j.cjco.2022.11.012

23. Vincent LT, Grant J, Ebner B, et al. Effect of Gender on Prognosis in Patients With Takotsubo Syndrome (from a Nationwide Perspective). Am J Cardiol. 2022;162:6–12. doi:10.1016/j.amjcard.2021.09.026

24. Waqar A, Jain A, Joseph C, et al. Cardioprotective Role of Estrogen in Takotsubo Cardiomyopathy. Cureus. 2022;14(3):e22845. Published 2022 Mar 4. doi:10.7759/cureus.22845

25. Fu L, Zhang H, Ong’achwa Machuki J, et al. GPER mediates estrogen cardioprotection against epinephrine-induced stress. J Endocrinol. 2021;249(3):209–222. Published 2021 May 20. doi:10.1530/JOE-20-0451

26. Shah RM, Shah M, Shah S, Li A, Jauhar S. Takotsubo Syndrome and COVID-19: Associations and Implications. Curr Probl Cardiol. 2021;46(3):100763. doi:10.1016/j.cpcardiol.2020.100763

